# Inference of causal and pleiotropic effects with multiple weak genetic instruments: application to effect of adiponectin on type 2 diabetes

**DOI:** 10.1101/2023.12.15.23300008

**Authors:** Paul M McKeigue, Andrii Iakovliev, Athina Spiliopoulou, Buddhiprabha Erabadda, Helen M Colhoun

## Abstract

Current methods for Mendelian randomization (MR) analyses are restricted to single SNP instruments, and cannot reliably infer causality with instruments that are mostly weak and pleiotropic. We describe methods to overcome these limitations: key innovations are construction of scalar instruments from multiple SNPs, use of a regularized horseshoe prior, and hypothesis tests based on the marginal likelihood of the causal effect parameter. To demonstrate the approach, we constructed genotypic instruments from unlinked *trans*-pQTLs detected in two large GWAS studies of plasma proteins, and tested the top 20 genes for which the aggregated effects of the instruments was associated with type 2 diabetes in the UK Biobank cohort. The only protein with clear evidence of a causal effect on type 2 diabetes was adiponectin, encoded by *ADIPOQ*: standardized log odds ratio -0.34 (95% CI -0.44 to -0.24) using UK Biobank instruments. These results have implications for the design and analysis of Mendelian randomization studies. Where the exposure under study is expression of a gene, restricting the instruments to *cis*-acting variants is likely to miss causal effects. Tests based on the marginal likelihood should supersede other methods of testing for causality in the presence of pleiotropy.

## Introduction

Instrumental variable analysis with genetic instruments (“Mendelian randomization”) has been widely used to infer causal effects of exposures (broadly defined to include behavioural traits, biomarkers and gene expression levels) on diseases. The biggest methodological challenge is how to infer causality when some of the genetic instruments have direct (pleiotropic) effects on the outcome that are not mediated through the exposure under study. These pleiotropic effects are usually not directly observed, and their distribution over the instruments is unknown. Inference of causality in the presence of pleiotropic effects has relied on makeshift procedures for the construction of estimators such as weighted medians that downweight or exclude instruments that are outliers.^1^ The results depend on which estimator is used: recent guidelines suggest that “investigators should pick a sensible range of methods to assess the sensitivity of their findings.”^2^

The motivation for the work reported here was to infer causality from genetic instruments constructed from *trans*-QTLs that perturb expression (as levels of the transcript or encoded protein) of a gene. Mendelian randomization studies of gene expression are usually restricted to *cis*-QTLs, on the basis that *trans*-effects are too weak and pleiotropic to be used as genetic instruments.^2^ Weak instruments are excluded because weak estimators constructed from ratios of estimated coefficients have unstable sampling properties where the denominators of these ratios – the instrument-exposure coefficients – can take values close to zero. With recently-described Bayesian methods that use an unregularized horseshoe prior to marginalize over the distribution of pleiotropic effects to infer the causal effect,^3,4^ there is no need to exclude weak instruments. However few real-world application of these methods have been reported.

For studies that use *trans*-QTLs as instruments, a key limitation of all existing methods is that they cannot aggregate the effects of multiple SNPs at each locus to construct unlinked instruments that can be modelled as independent. The usual procedure is to select a single SNP that is reliably associated with the exposure from each genomic region in which genetic associations with the exposure have beeen detected. This does not use all the available information about the effects of variants in the region on the exposure, and the results may depend upon which variants are selected.

This paper describes methods that overcome these limitations, and demonstrates their application to infer causal effects of circulating proteins on type 2 diabetes in the UK Biobank cohort.

## Methods

### Statistical model

For a Mendelian randomization study with *J* unlinked genetic instruments, we specify a model with three parameters:

- *α* vector of coefficients of effects of the instruments on exposure *X*.
- *β* vector of coefficients of direct (pleiotropic) effects of the instruments on outcome *Y*
- *θ* causal effect of *X* on *Y*

From summary statistics we have estimates 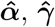 of the effects *α, γ* of the *J* instruments on the exposure and the outcome, with corresponding standard errors *s*_*α*_, *s*_*γ*_. We assume that these estimates have been adjusted for relevant covariates such as genetic background. As the instruments are unlinked, we can model the estimates as independent Gaussian variables conditional on the true values.

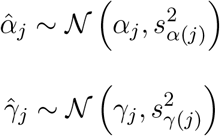

The crude effect of each instrument on the outcome is the sum of the direct effect and the causal effect.

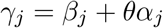

With priors on *β, θ* we can compute the joint posterior distribution of these parameters given 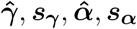. As the form of the distribution of pleiotropic effects over loci is unknown, any realistic statistical model has to specify a prior on these effects that encompassses a broad family of symmetric distributions ranging from a spike-and-slab mixture to a Gaussian. One such prior is the horseshoe,^5^ which has been applied to inference of causal effects from Mendelian randomization using individual-level data^3^ or summary-level data.^4^ A limitation of the original horseshoe is that the posterior distribution is difficult to sample from because the curvature varies. This problem is not obvious when Gibbs sampling is used,^4^ but is revealed by the divergence diagnostics that are implemented in the NUTS (No U-Turn Sampling) algorithm used in Stan and other probabilistic programming languages.^6^ Another limitation of the original horseshoe is that there is no way of specifying a realistic prior on the size of the nonzero effects in the “slab” component. When modelling the genetics of complex traits, we already know that the effects of common genetic variants on a complex trait are usually small. The regularized horseshoe prior, known as the “Finnish horseshoe”, overcomes these limitations as described below. We name our method “MR-Hevo”, using the Finnish word for a horse to distinguish it from the recently-described “MR-Horse” method which uses an unregularized horseshoe prior.^4^

Fig 1 shows the model as a directed acyclic graph in plate notation. The regularized horseshoe prior for the regression coefficients *β*_1_, …, *β*_*J*_ is

**Fig 1.**
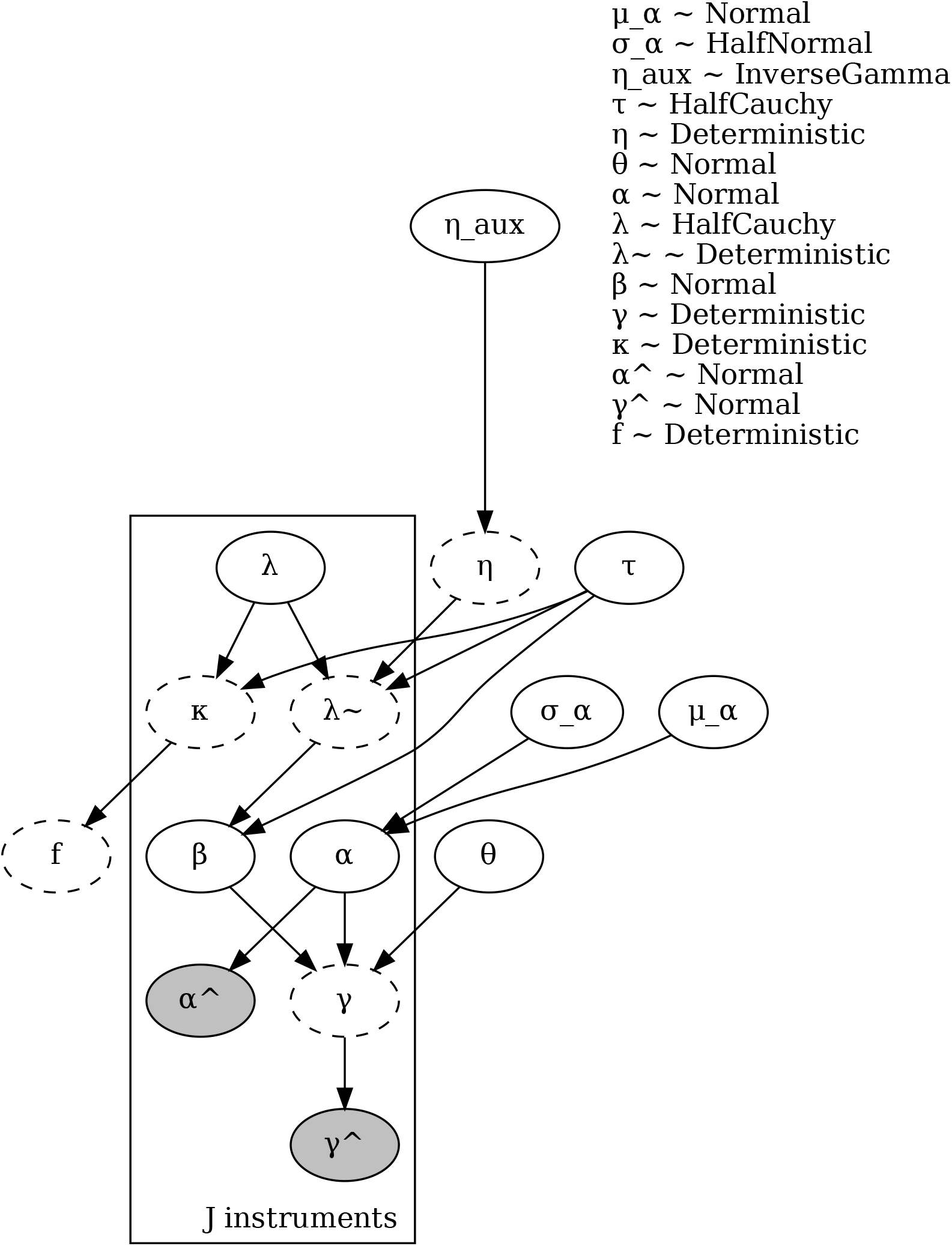
MR-Hevo model as a directed acyclic graph in plate notation. Stochastic nodes are shown as ellipses with continuous borders, deterministic nodes as ellipses with dashed borders. Observed nodes are shaded.

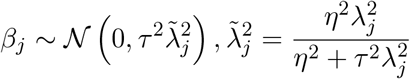

Half-Cauchy priors are specified on the unregularized local scale parameters *λ*_*j*_ and the global scale parameter *τ*.

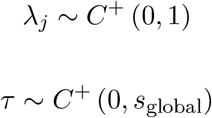

The heavy tail of the half-Cauchy priors on *λ*_*j*_ allows some of the regression coefficients to escape the shrinkage imposed by small values of the global parameter *τ*. These nonzero coefficients are the slab component of the spike-and-slab distribution.

A weakly informative prior is specified for the regularization parameter *η*:

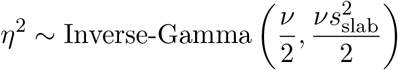

Even the largest direct effects will be regularized as a Gaussian with standard deviation *η*. The scaling factor *s*_slab_ is specified based on prior information about the expected size of the largest direct effects. To constrain *η* so that the sampling algorithm does not diverge, we set ν = 2; this translates to a half-*t*_2df_ prior on the scale of the largest direct effects.

### Shrinkage coefficients and effective number of nonzero effects

Calculation of the shrinkage coefficients and the effective number of nonzero parameters does not affect the results of statistical modelling, but is helpful for interpretation of what the horseshoe prior is learning.

The shrinkage coefficient *κ*_*j*_ for the *j*th regression coefficient *β*_*j*_, with prior 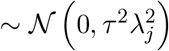 and Gaussian likelihood with Fisher information ℐ_*j*_ can be defined as the fraction by which the information in the prior shrinks the posterior mean of *β*_*j*_ from the maximum likelihood estimate:

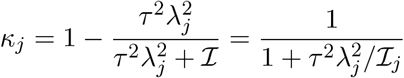

This definition of the shrinkage coefficient ignores the regularization parameter *η*, which imposes a lower bound on the shrinkage of *β*_*j*_. The prior on each shrinkage coefficient has a horseshoe shape. The shrinkage coefficients *κ*_*j*_ can take values from 0 (no shrinkage) to 1 (complete shrinkage).

ℐ_*j*_ is asymptotically equivalent to the inverse variance of the maximum likelihood estimate of *β*_*j*_ at *θ* = 0. When using summary statistics, the approximate variance of the maximum likelihood estimate of the ratio *β*_*j*_ = *γ*_*j*_*/α*_*j*_ can be calculated by the delta method. The effective fraction *f* of nonzero coefficients is

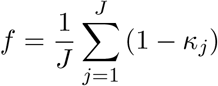

The prior expectation of *f* is

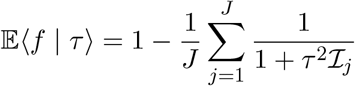

The information ℐ_*j*_ on the coefficient *β*_*j*_ depends on the sample sizes in the studies from which the summary statistics 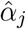 and 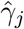 were obtained. With more information, a smaller value of *τ* is required to impose the same prior expectation of *f*. Piironen and Vehtari recommend setting the scale of the prior on *τ* to be consistent with this prior expectation.^6^ For this analysis we have set the scale of the Cauchy prior on *τ* to so that the prior median *τ*_0_ is the value at which the prior expectation of *f* given ℐ_1_, …, ℐ_*J*_ is 0.2, and we have set the scale of the Inverse-Gamma prior on *η*^2^ as 0.1, encoding our prior knowledge that effects of any single genomic region on type 2 diabetes are of modest size. We examined the sensitivity of the results to these prior settings as described below.

### Computational methods

The NUTS algorithsm is implemented in several recently developed probabilistic programming languages, including Stan, PyMC, and NumPyro. MR-Hevo is implemented in NumPyro, which has a more robust implementation of the NUTS algorithm than Stan giving fewer divergent transitions. To improve posterior geometry, the Cauchy distributions are parameterized as mixture distributions (Gaussian with inverse gamma distribution of scale parameter). A divergent transitions rate of less than 1 in 1000 is considered acceptable. This criterion was met with the regularized horseshoe, but not with the original horseshoe for which the divergent transition rate was about 5%. To obtain the marginal likelihood as a function of the parameter of interest, we simply divide the posterior by the prior on that parameter. We calculated the likelihood of the causal effect parameter *θ* by fitting a kernel density to the posterior samples of *θ*, weighting each observation by the inverse of the prior. A quadratic function was fitted by least-squares to the logarithm of this likelihood function. Where the log-likelihood is asymptotically quadratic, a confidence interval for *θ* and a test of the null hypothesis *θ* = 0 can be obtained by standard methods. Where the log-likelihood is not quadratic, inference should be based on the posterior distribution rather than on sampling theory.

### Constructing scalar instruments from multiple SNPs

#### Instrument-exposure coefficients

From summary statistics we have for each clump of exposure-associated SNPs a vector 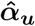 of univariate coefficient estimates for the effect of the SNPs on the exposure. Multivariable coefficient estimates 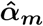 are calculated by premultiplying the univariate coefficients by the inverse of the correlation matrix **Σ**_***G***_ between the SNP genotypes.

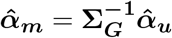

The correlation matrix **Σ**_***G***_ is obtained from a reference panel such as 1000 Genomes. A shrinkage penalty (equivalent to ridge regression) can be imposed by adding a penalty factor to the diagonal elements of **Σ**_***G***_. Where the correlation matrix is singular or ill-conditioned, a pseudo-inverse can be used to calculate the multivariable coefficients. For an individual with genotypes ***G*** at the exposure-associated SNPs, a locus-specific score *S* predicting the exposure is calculated as 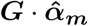.

Because the score *S* is calculated from the genotypes and the genotype-exposure coefficients, we cannot use it as a genotypic instrument in a model of the relationship of the genotype-outcome coefficients to the genotype-exposure coefficients. We can factor the dot product 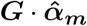 as the product of two scalars: the magnitude of the multivariable coefficient vector 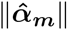 and a pseudo-genotype 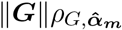, where 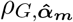 is the correlation between ***G*** and 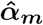, geometrically equivalent to the cosine of the angles between these vectors. We can then substitute 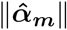 for the scalar coefficient estimate 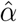 and 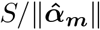 for the scalar instrument *Z* as a pseudo-genotype in the statistical model. The derived coefficient for the estimated effect of the instrument on the exposure is always positive; flipping the coding of the alleles would flip the sign of *ρ*_*G,α*_. We can calculate the standard error of 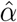as 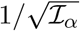 where ℐ_*α*_ is the Fisher information on the slope of the linear regression of *X* on *Z*, given by

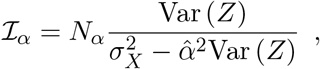

where *N*_*α*_ is the sample size of the study from which the estimated univariate coefficients 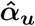 were obtained, Var (*Z*) is the variance of the scalar instrument *Z* estimated in a reference panel, and 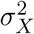 is the variance of the exposure *X*. If an estimate of 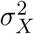 is not given, it can be calculated from the allele frequency *a*_*j*_ of the *j*th SNP, the standard error *s*_*G*(*j*)_ of the *j*th univariate coefficient estimate 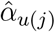, and the sample size *N*_*α*_ as

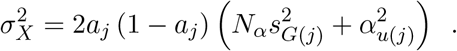

#### Instrument-outcome coefficients

For this study, where we have individual-level data in UK Biobank for the scalar instrument *Z* and the outcome *Y*, we estimated the coefficients for the effect of the scalar instruments *Z* on the outcome *Y* and its standard error by fitting logistic regression models. A method for calculating the instrument-outcome coefficient 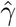 and its standard error from summary-level data for SNP-outcome coefficients is described below.

For each instrument we standardize the pseudo-genotypes *Z* to unit variance and scale the coefficient estimates accordingly, so that the variance explained by the *j*th linear predictor 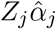, which can be interpreted as the “strength of the instrument”, is proportional to the square of the scalar coefficient 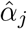. The prior on the scale of the pleiotropic effects *β* of the instruments *Z* is thus independent of the strength of the instrument.

Where only summary-level data are available for the genotype-outcome associations, an estimate 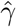 of the coefficient for the effect of the scalar instrument *Z* on the outcome and its standard error can be obtained using individual-level genotype data from a reference panel. First, the vector 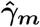 of estimated multivariable coefficients is calculated by premultiplying the estimates 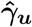 of the univariate coefficients by 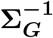. The coefficient 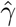 for the scalar instrument *Z* is then estimated by minimizing the sum of the squared differences between the predictor calculated from *Z* and the linear predictor calculated from the vector of genotypes ***G***, where the sum is taken over the genotypes of all *n* individuals in the reference panel.

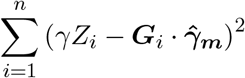

Equating to zero the derivative of this expression with respect to *γ*, substituting 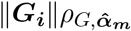 for *Z*_*i*_ and factoring the dot product 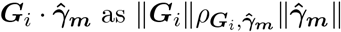 we obtain

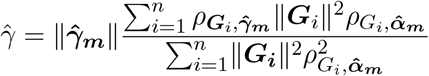

The ratio in this expression can be recognized as a weighted average of the ratio 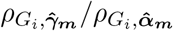, with weights 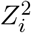. The standard error of 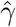 is 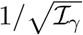 where ℐ_*γ*_ is the Fisher information on the slope of the regression of *Y* on *Z*, given (for a logistic regression model) by

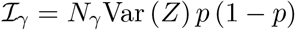

where *N*_*γ*_ and *p* are respectively the total sample size and the the proportion of cases in the dataset from which the coefficient estimates 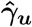 were obtained.

### Example: effect of adiponectin on type 2 diabetes

As the methods described above overcome some of the limitations of standard methods for design and analysis of Mendelian randomization studies, they may help to resolve contradictory results obtained in earlier studies. As an example, we examine the effect of adiponectin, encoded by the *ADIPOQ* gene, on the risk of type 2 diabetes. Low adiponectin levels are strongly associated with type 2 diabetes. Of four previous studies using Mendelian randomization to investigate whether low adiponectin levels cause type 2 diabetes, two reported no evidence of a causal effect^7,8^ and two reported evidence for a causal effect.^9,10^ We obtained coefficient estimates for the effects of SNPs on circulating levels of adiponectin from two studies:

- The DeCODE study of 35559 Icelanders in which 4719 proteins on the SomaLogic v4 panel were measured in plasma.^11^ 2207 aptamers on this platform that appeared to cross-react with complement factor H were excluded. The criteria for identifying these aptamers were: a *trans*-pQTL at the *CFH* locus and no *cis*-pQTL for the protein that the aptamer was designed to detect and association of the *trans*-score with age-related macular degeneration.
- The UK Biobank proteomics study of 54306 individuals in which 2923 proteins on the Olink Explore panel were measured in plasma^12^

The calculated standard deviation of adiponectin levels was 1.14 in the DeCODE dataset and 0.83 in the UK Biobank dataset. In each genomic region where there was at least one SNP-exposure association with *p* < 10^*−*6^, all SNPs with *p* < 10^*−*6^ separated by no more than 1 Mb were included in the calculation of the multivariable coefficient vector. This yielded 26 unlinked genotypic instruments from the DeCODE study, calculated from 567 SNPs, and 44 instruments from the UK Biobank proteomics study, calculated from 945 SNPs. Each instrument was annotated with a list of nearby genes, and the genes on each list were matched to a list of 256 genes for which SNP associations with type 2 diabetes have been reported at *p* < 5× 10^*−*8^, extracted from GWAS Catalog.

The target dataset with genotypes and type 2 diabetes status was the UK Biobank cohort. Ethical approval for the UK Biobank study was granted in 2011 by the North West Multi-centre Research Ethics Committee (11/NW/0382), and renewed every five years since then. Informed consent was obtained for all participants in UK Biobank. The work described herein was approved by the UK Biobank under application number 23652.

Separate analyses were undertaken using the instruments constructed from these two proteomics studies. When using instruments obtained from the UK Biobank proteomics study, the 54306 participants who were in the proteomics study were excluded from the target dataset.

Because the individuals in the proteomics studies were predominantly of European ancestry, the target dataset was restricted to individuals of European ancestry. Of 451447 unrelated individuals with nonmissing phenotype and genotype data, 37981 were classified as cases of type 2 diabetes and the all others were classified as noncases.

For each instrument, the pseudo-genotypes were standardized to unit variance and a logistic regression model was fitted with type 2 diabetes as outcome variable. The other covariates were sex and the first five principal components of the SNP relationship matrix. The coefficients for the effect of each scalar instrument on type 2 diabetes were used to fit the model. *Cis*-pQTLs were excluded.

For comparison with inference based on the marginal likelihood of the causal effect parameter, three widely-used estimators for the causal effect were calculated from the coefficient ratios: the inverse-variance weighted mean of the coefficient ratios which assumes no pleiotropy, the weighted median, and the penalized weighted median which downweights outliers.^1^ The coefficient ratio estimates and their standard errors were calculated by the delta method, based on second-order Taylor expansions for the moments of the distribution of the ratio of two independent normally-distributed variables. Standard errors for the weighted median and penalized weighted median estimators were calculated by a parametric bootstrap method. To generate the sampling distribution of the weighted median, we used 4000 draws from the posterior distribution of the regularized shrinkage parameters 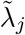 and the instrument effects *α*_*j*_. At each draw we sampled the direct effects *β*_*j*_ of *J* new genetic instruments, and sampled the coefficient ratio estimates 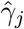 (calculated by the delta method) conditional on *β*_*j*_, *α*_*j*_ and the standard errors s 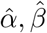 of these estimates. The standard deviation of the weighted median of the coefficient ratio estimates obtained from this posterior predictive distribution was calculated and used to obtain *p*-values and confidence intervals.

## Results

Table S1 shows summary statistics for the genetic instruments for adiponectin derived from each pQTL study. There is suprisingly little overlap between the 24 *trans*-pQTLs detected in the DeCODE study and the 43 *trans*-pQTLs detected in UK Biobank. Only eleven *trans*-pQTLs – gene-poor regions at 219.4 Mb on chromosome 1, 34.2 Mb on chr 6, 139.5 Mb on chromosome 6, and regions containing *ALAS1, ADRB1, SPON1, PDE3A, ABCB9, CDH13, AKR1B1P7*, and *MIR6813* – were detected in both studies at a threshold of *p* < 10^*−*6^. This may reflect the low power to detect weak *trans*-pQTLs even in these large datasets. In both studies there is a *cis*-QTL at the *ADIPOQ* transcription site. In the DeCODE study an extended *cis*-pQTL is detected 2 Mb upstream of the transcription site. Only two pQTLs – *SPON1* and *APOC1* – contained genes that were listed in GWAS Catalog as associated with type 2 diabetes.

Figure 2 shows that 22 of 24 *trans*-pQTL instruments from DeCODE and 39 of 43 *trans*-pQTL instruments from UKBB are inversely associated with type 2 diabetes. The *cis*-pQTL instruments are not associated with type 2 diabetes.

**Fig 2.**
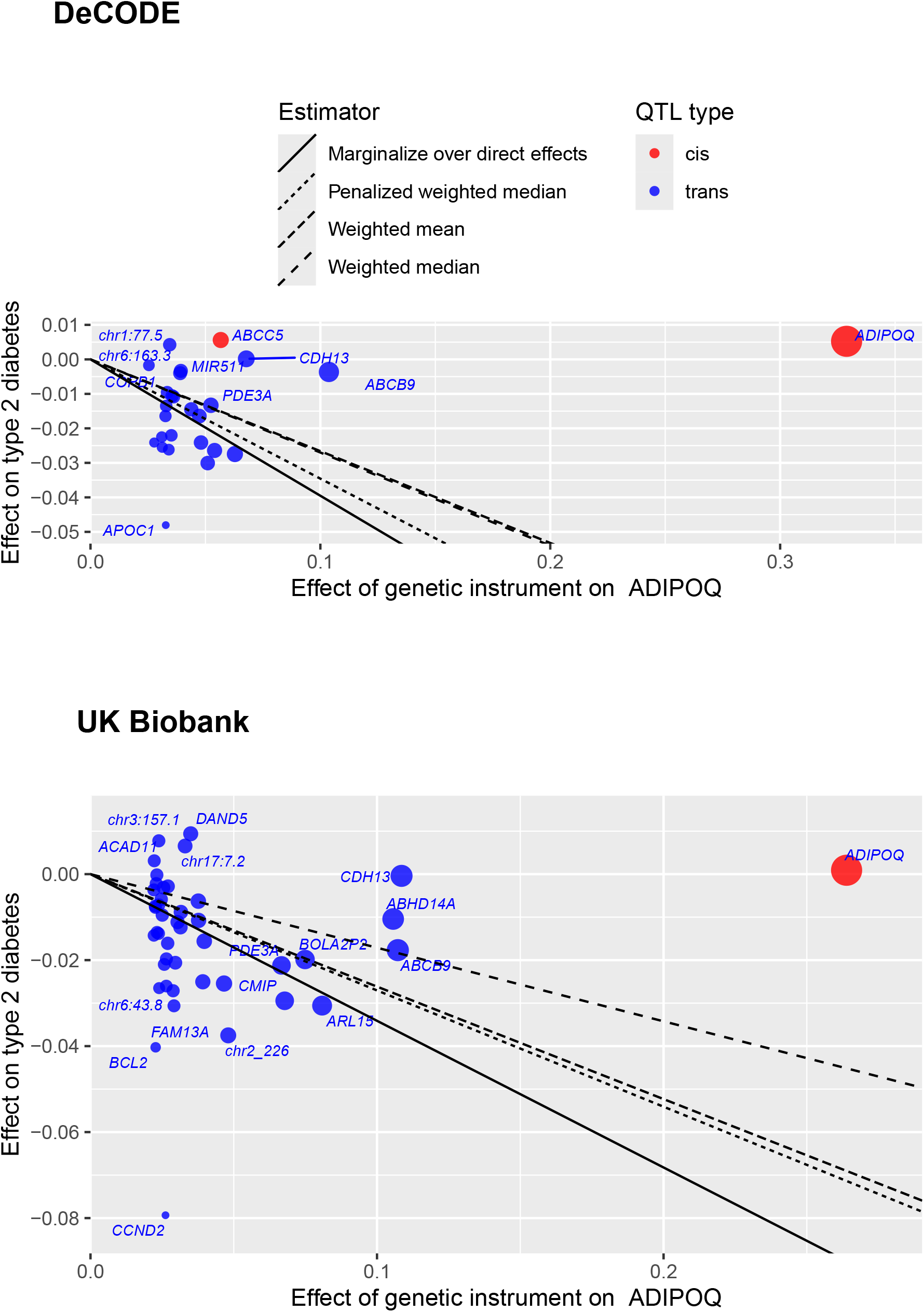
Plot of coefficients of regression of type 2 diabetes on each instrument against coefficients of regression of adiponectin levels on each instrument. Size of each data point is inversely proportional to the standard error of the ratio estimate. The value of each estimator is shown as the slope of a line passing through the origin. *Cis*-pQTLs are excluded from these estimates.

Table 2 shows the estimates of the causal effect parameter obtained by each method, as log odds ratios for unit change in adiponectin levels. The maximum likelihood estimates obtained by marginalizing over direct effects were -0.4 (95% CI -0.52 to -0.27) using DeCODE instruments and -0.34 (95% CI -0.44 to -0.24) using UK Biobank instruments. In comparison with the weighted median or penalized weighted median estimators, the confidence intervals obtained from the likelihood function were wider and the *p*-values were more conservative.

**Table 1.** Posterior summaries of global parameters of MR-Hevo models of the effect of instruments for adiponectin levels on type 2 diabetes.

| Parameter | Prior median | Posterior distribution |  |  |
| --- | --- | --- | --- | --- |
|  |  | Median | 10th centile | 90th centile |
| **DeCODE** |  |  |  |  |
| Global shrinkage $\tau$ | 0.07 | 0.008 | 0.004 | 0.014 |
| Regularization $\eta$ | 0.10 | 0.099 | 0.063 | 0.189 |
| Fraction nonzero pleiotropic effects $f$ | 0.20 | 0.036 | 0.014 | 0.078 |
| Causal effect $\theta$ | 0.00 | -0.399 | -0.470 | -0.326 |
| **UK Biobank** |  |  |  |  |
| Global shrinkage $\tau$ | 0.08 | 0.008 | 0.005 | 0.013 |
| Regularization $\eta$ | 0.10 | 0.091 | 0.059 | 0.170 |
| Fraction nonzero pleiotropic effects $f$ | 0.20 | 0.033 | 0.015 | 0.063 |
| Causal effect $\theta$ | 0.00 | -0.340 | -0.404 | -0.271 |

**Table 2.** Effect of adiponectin on type 2 diabetes: comparison of causal effect parameter estimates obtained with different methods.

| Estimator | Estimate | 95% CI | *p*-value |
| --- | --- | --- | --- |
| **DeCODE instruments** |  |  |  |
| Weighted mean | -0.27 | -0.32, -0.22 | $5 \times 10^{-25}$ |
| Weighted median | -0.27 | -0.47, -0.07 | 0.009 |
| Penalized weighted median | -0.35 | -0.47, -0.22 | $3 \times 10^{-8}$ |
| Marginal likelihood | -0.40 | -0.52, -0.27 | $2 \times 10^{-9}$ |
| **UK Biobank instruments** |  |  |  |
| Weighted mean | -0.26 | -0.3, -0.22 | $2 \times 10^{-37}$ |
| Weighted median | -0.17 | -0.3, -0.04 | 0.01 |
| Penalized weighted median | -0.27 | -0.4, -0.14 | $3 \times 10^{-5}$ |
| Marginal likelihood | -0.34 | -0.44, -0.24 | $1 \times 10^{-10}$ |

Supplementary Figure S2 shows the posterior distribution and log-likelihood of the causal effect parameter using UK Biobank instruments. As expected with this large sample size, the posterior distribution is approximately Gaussian and the log-likelihood is well approximated by a quadratic function. Supplementary Table 1 shows posterior summaries of the model parameters. For each of the two sets of instruments, the prior median of the global shrinkage parameter *τ* was set to a value that translated to an expectation of 0.2 for the effective fraction *f* of instruments with nonzero pleiotropic effects. The posterior medians of *τ* and *f* were much lower than the prior medians. Consistent with this low estimate of the fraction of instruments that have nonzero pleiotropic effects, Figure 3 shows that only a few instruments have clearly escaped shrinkage: *of the DeCODE instruments these were APOC1 CDH13* and *ABCB9*, and of the UK Biobank instruments these were *CDH13, ABCB9* and *CCND2*.

**Fig 3:**
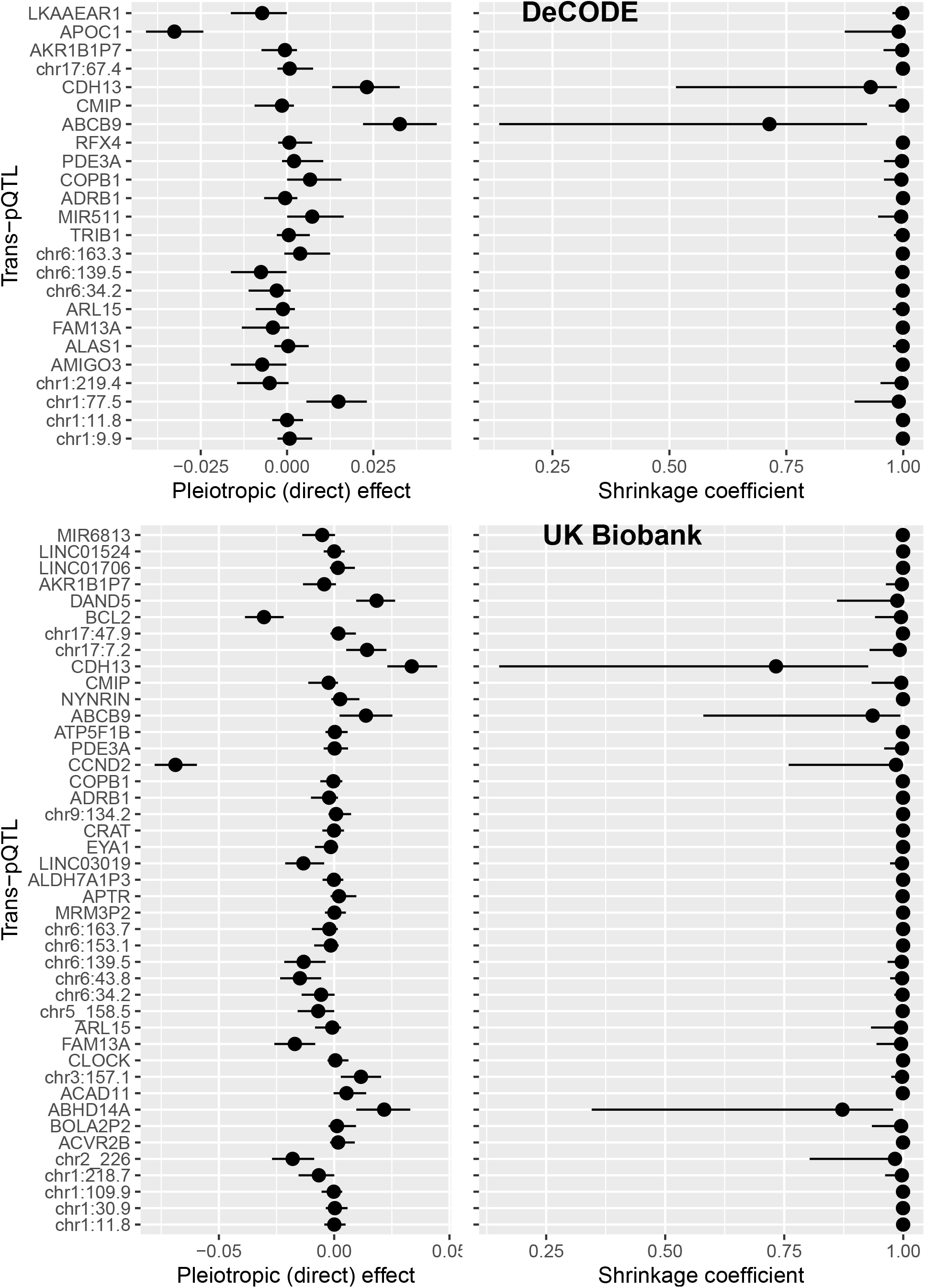
Effects of trans-QTLs for adiponectin on type 2 diabetes: posterior medians and 80% credible intervals for direct effects and shrinkage coefficients

Specifying different values for the scale of the prior on the global shrinkage parameter *τ* which regulates the number of nonzero pleiotropic effects, or for the scale of the prior on the regularization parameter *η* which regulates the size of these effects, did not appreciably alter the results.

Fig S3 compares, for the top 20 genome-wide aggregated *trans*-effect scores associated with type 2 diabetes in the UK Biobank cohort, the *p*-values calculated from the weighted median and penalized weighted median estimators (using the procedure described above to calculate the standard deviation of these estimators), with *p*-values based on the marginal likelihood. *ADIPOQ* was the only gene for which *p*-values based on the marginal likelihood provided strong evidence of a causal effect. The p-values obtained from tests based on the sampling distribution of the weighted median were broadly concordant with those obtained from tests based on the marginal likelihood but were less extreme. This is to be expected, as the median is less efficient than the maximum likelihood estimate.

## Discussion

Mendelian randomization analysis, combined with related approaches including transcriptom-wide association studies and co-localization analysis, has been widely used to infer causal effects of gene expression on outcomes. The most widely-used methods use summary statistics for the coefficients for the effect of each SNP on the outcome and on the exposure from non-overlapping samples.^13^ “Estimators” for the causal effect are constructed from the ratios of SNP-outcome to SNP-exposure coefficients. Bayesian methods for marginalizing over the unobserved pleiotropic effects to infer the causal effect parameter in a Mendelian randomization study have been described previously.^3,4^ The methods described in this paper extend this approach: (1) by allowing unlinked scalar genetic instruments to be constructed from all SNPs associated with the exposure without having to restrict to one SNP from each genomic region; (2) by using a regularized horseshoe prior which allows encoding of prior knowledge that genetic effects of a single genomic region on a complex trait are usually small; and (3) by constructing classical hypothesis tests based on the likelihood of the causal effect parameter.

The difficulties associated with using ratios of coefficients to construct estimators are eliminated in a Bayesian framework, where nothing is “estimated”. Where the log-likelihood is asymptotically quadratic, statistical theory guarantees that the maximum-likelihood estimate has the sampling properties that are desirable for an estimator: consistency, minimum variance and unbiasedness. Where the log-likelihood is not approximately quadratic, it is preferable to base inference directly on the likelihood function rather than on the sampling properties of an estimator. Because Bayesian inference does not rely on the sampling properties of ratios, there is no need to exclude “weak instruments”. This makes it possible to integrate Mendelian randomization into genome-wide aggregated *trans*-effects analysis, which attempts to identify core genes on which the polygenic effects of many variants coalesce via *trans*-effects to influence complex traits.^14^ In this approach, the genetic instruments are clumps of SNPs with *trans*-effects on the expression of a gene as transcript or circulating protein.

Using instruments constructed from *trans*-pQTLs in two different studies that used different assays for the protein, there is clear support for a causal effect of low adiponectin levels on type 2 diabetes. This is apparent even without a formal statistical analysis, as the signs of the effects of the instruments are almost all negative. Earlier studies that failed to detect a causal effect were restricted to *cis*-acting SNPs.^7,8^ Some of these *cis*-acting SNPs affect not only the measured levels but also the splicing of the gene product,^15^ so their effects on measured adiponectin levels do not necessarily correspond to effects on the functional protein. The most direct human evidence for a causal effect of low adiponectin on type 2 diabetes comes from reports of families in which type 2 diabetes segregated with rare loss-of-function variants in *ADIPOQ*.^16^ In the obese mouse model, overexpression of adiponectin in the liver protected against diabetes.^17^ Though adiponectin analogues have been studied experimentally, none has reached the clinical stage of drug development. Misleading results from *cis*-Mendelian randomization studies may have discouraged development of a possible drug target.

Where, as in this example, the exposure under study is a gene transcript or a circulating protein, study designs that use only *cis*-acting variants as genetic instruments have been advocated.^18^ The rationale for thse “*cis*-MR” designs is that *cis*-acting variants are less likely to have pleiotropic effects on the outcome that are not mediated through the level of the circulating protein. This supposition is questionable: especially where *cis*-acting variants affect the splicing of the transcript, the relationship between measured levels and functional activity of the protein may be broken. More fundamentally, restriction to *cis*-QTLs focuses on the genes least relevant to disease risk. Most disease/trait-associated SNPs are not co-localized with *cis*-eQTLs detected in relevant cell types.^19^ Recent studies have shown that disease-relevant genes are enriched with redundant enhancer domains and depleted of *cis*-eQTLs of large effect, as would be expected if perturbation of these genes has large effects on fitness.^20,21^ Most of the SNP heritability of gene expression is attributable to *trans*-effects.^22^

Bayesian hypothesis testing is based on the Likelihood Principle: all information conveyed by observations that supports one model or one parameter value over another is contained in the ratio of the likelihoods of these models or parameter values. The likelihood-based approach to causal inference described in this paper is aligned with the not yet universally-accepted principle that causal inference is just a special case of statistical inference, requiring attention to assumptions about exchangeability between observations (of exposure and outcome) and predictions of the effect of perturbing the exposure.^23^ In this decision-theoretic framework there is no need to invoke counterfactuals^24^ or to require causal effects to be expressed as marginal rather than as conditional effects. In this example, the likelihood of a model with a causal effect parameter and only a few nonzero pleiotropic effects is higher than the likelihood of a model with no causal effect that requires more nonzero pleiotropic effect parameters to adjust to the data to obtain the same fit. Thus even though the causal effect parameter is not identifiable because for any setting of the causal effect parameter we can find a setting of pleiotropic effect parameters that has the same fit to the data, a formal hypothesis test can be obtained. With horseshoe priors, which allow the distribution of the pleiotropic effects to be learned from the data, this model comparison is performed “under the hood,”^25^ without the computational inconvenience of having to average over all possible partitions of the variables into two disjoint sets (spike-and-slab) as in the contamination mixture model^26^ or over different settings of the number of nonzero effect parameters.^27^ A corollary of inference based on model comparison is that reliable inference of causality in the presence of pleiotropy requires a relatively large number of unlinked genetic instruments to allow learning the distribution of pleiotropic effects.

The statistical model assumes that the effects of the instruments on the exposure and their direct effects on the outcome are independent in magnitude and direction: this has been denoted the InSIDE (Instrument Strength Independent of Direct Effect) assumption^28^ though this term has also been used for the less stringent assumption that “the magnitude of the pleiotropic effects are independent of the SNP-exposure associations.”^29^ With the procedure described here for constructing scalar instruments, the estimates for the coefficients of the instrument-exposure effects can take only positive values (a negative effect would simply correspond to flipping of the allele coding), and the direct effects are assumed to arise from a distribution with zero mean. This assumption that the instrument-exposure effects and the direct effects are uncoupled in direction is termed “balanced pleiotropy”. It is possible in principle to infer a causal effect if the instrument-exposure effects and the direct effects are assumed to be uncoupled in magnitude even if they are coupled in direction^28^ but it is not obvious why this would be a biologically realistic assumption.

If the statistical model is extended to allow the instrument-exposure effects and the direct effects to be coupled in magnitude and direction, it is not possible to infer causality without other information about pleiotropy because models with and without a causal effect can fit the data equally well with the same number of adjustable parameters. It may be more useful to formulate and test possible hypotheses about mechanisms that could give rise to coupling of instrument-exposure effects with direct effects on the outcome, as this may identify a shared aetiological pathway. For adiponectin and type 2 diabetes, for instance, coupling could arise if the pQTLs for adiponectin are QTLs for a trait such as obesity that causes low adiponectin levels and increased risk of type 2 diabetes. In this situation other circulating proteins such as leptin that are biomarkers for obesity would show a similar pattern of apparently causal pQTL effects on type 2 diabetes. More generally, pQTLs that have pleiotropic effects should be detectable on a heat map of correlations between genotypic scores for multiple proteins or transcripts.

Guidelines for the design of Mendelian randomization studies have recently been updated.^2^ Based on the work described here, some suggestions for revisions to these guidelines are listed below.

- For inference of causal effects where pleiotropic effects may be present, the optimal method is to compute the likelihood of the causal effect parameter by marginalizing over the distribution of pleiotropic effects. This is robust to the inclusion of weak instruments, and is computationally efficient with modern tools for Bayesian computation. Investigators should not “pick a range of methods”, but assess the sensitivity of their results to prior assumptions about the distribution of unobserved pleiotropic effects. There is no advantage to using tests based on the sampling distribution of estimators such as the weighted median, when the likelihood can be calculated as a function of the parameter of interest.
- There is no need to restrict the selection of SNPs to one SNP from each genomic region: the methods described here allow scalar instruments to be constructed from multiple SNPs. This maximize the strength of the genetic instruments that can be obtained for each genomic region, and ensures that the results do not depend upon an arbitrary choice of which SNP to select from each region.
- Study designs that rely on using variants from a single genomic region, for instance using only *cis*-acting variants to study the effects of a gene transcript or gene product on a disease or trait, are likely to give misleading results and should be deprecated. Because *cis*-acting variants are likely to affect the function of the gene through mechanisms (such as altered splicing) other than the measured levels of the transcript or gene product, it is preferable to report the effects of a *cis*-QTL separately.

## Data Availability

UK Biobank data are available to approved researchers via managed access. A guide to accessing data is available at https://biobank.ctsu.ox.ac.uk/crystal/exinfo.cgi?src=accessing_data_guide

https://github.com/molepi-precmed/mrhevo.

## Declarations

### Data and code availability

A Python script using NumPyro to fit the MR-Hevo model, together with the summary-level data used in this paper, is available at https://github.com/molepi-precmed/mrhevo.

## Acknowledgements

No specific funding was received for this work. This research has been conducted using the UK Biobank Resource under application number 23652. The development of the GENOSCORES platform was supported by a Springboard Award (SBF006/1109) to AS from the Academy of Medical Sciences, supported in turn by the Wellcome Trust, the UK Government Department of Business, Energy and Industrial Strategy, the British Heart Foundation, and Diabetes UK. AI was supported by the Medical Research Council Cross Disciplinary Fellowship (XDF) Programme (MC_FE_00035). HC is supported by an endowed chair from the AXA Research Foundation.

## Web resources

GWAS Catalog: https://www.ebi.ac.uk/gwas/

## Supplementary information

### Supplementary Figures

**Fig S1.**
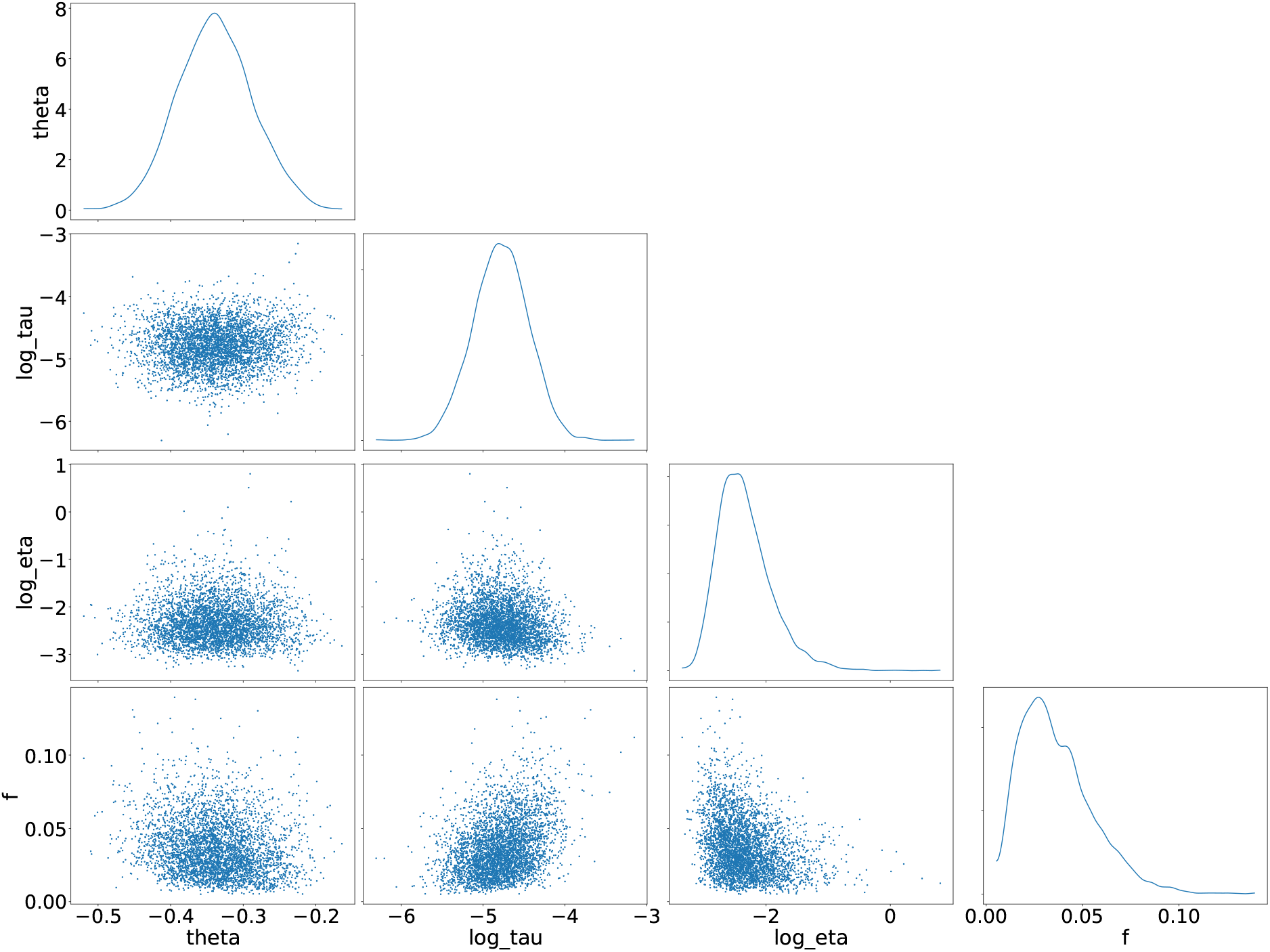
Pairs plot of posterior samples using UK Biobank instruments for adiponectin levels

**Fig S2.**
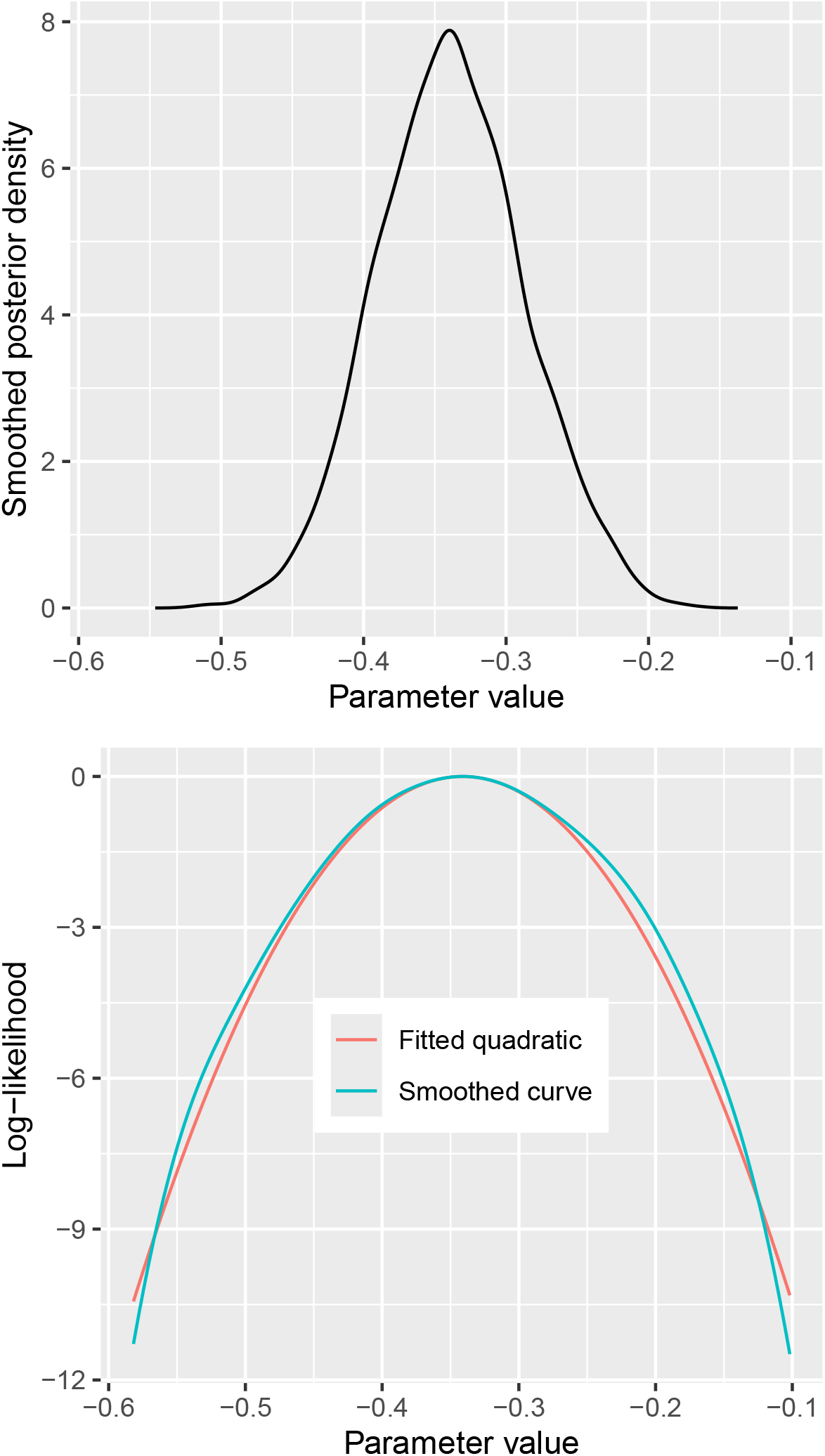
Posterior density and log-likelihood of causal effect parameter *θ* for effect of adiponectin on type 2 diabetes, using UK Biobank instruments

**Fig S3.**
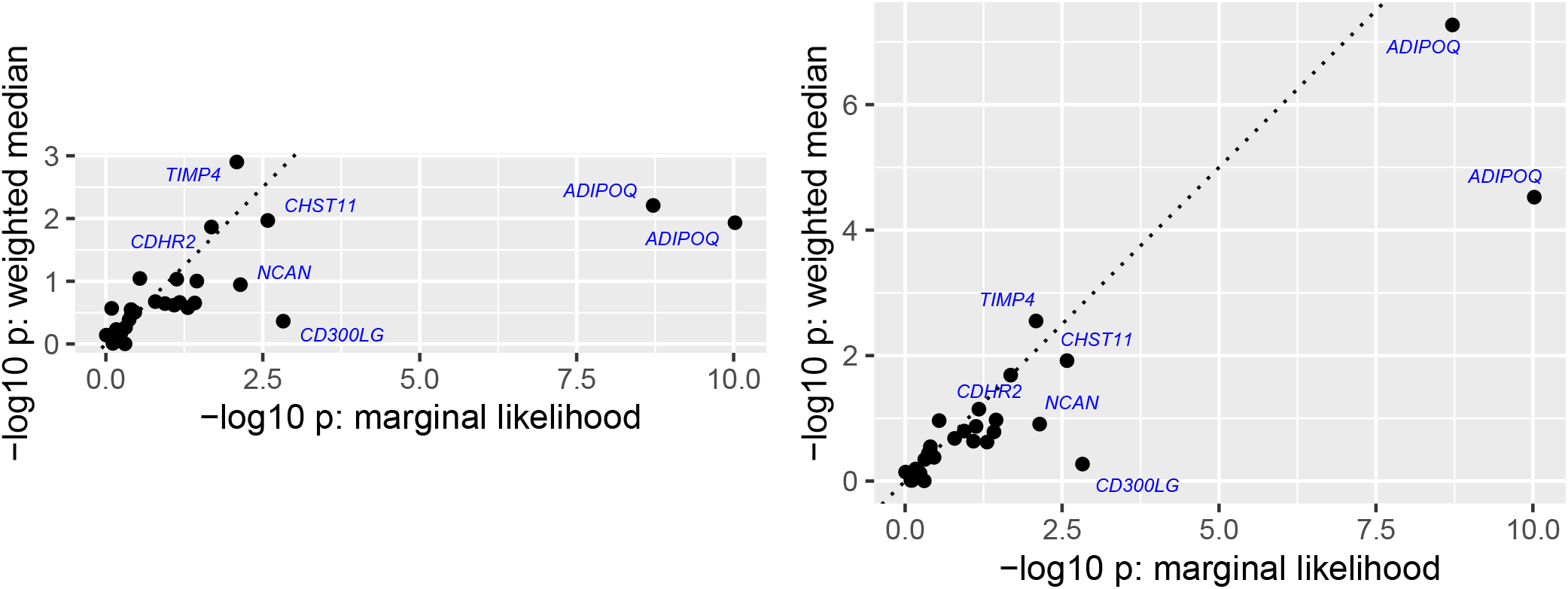
Comparison of p-values from weighted median estimators with p-values from marginal likelihood, for top 20 associations of genome-wide aggregated *trans*-scores with type 2 diabetes in UK Biobank cohort

### Supplementary Tables

**Table S1.**
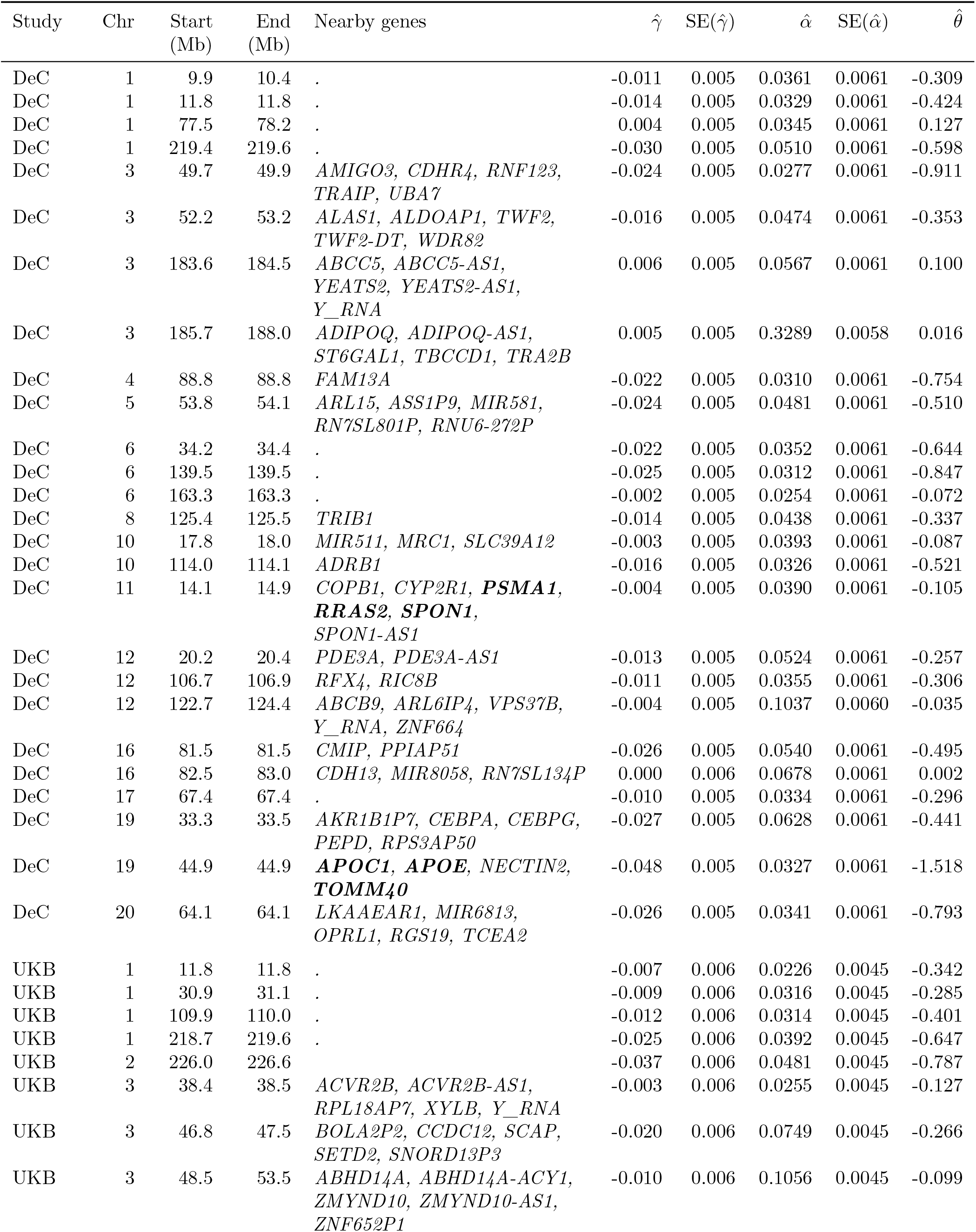
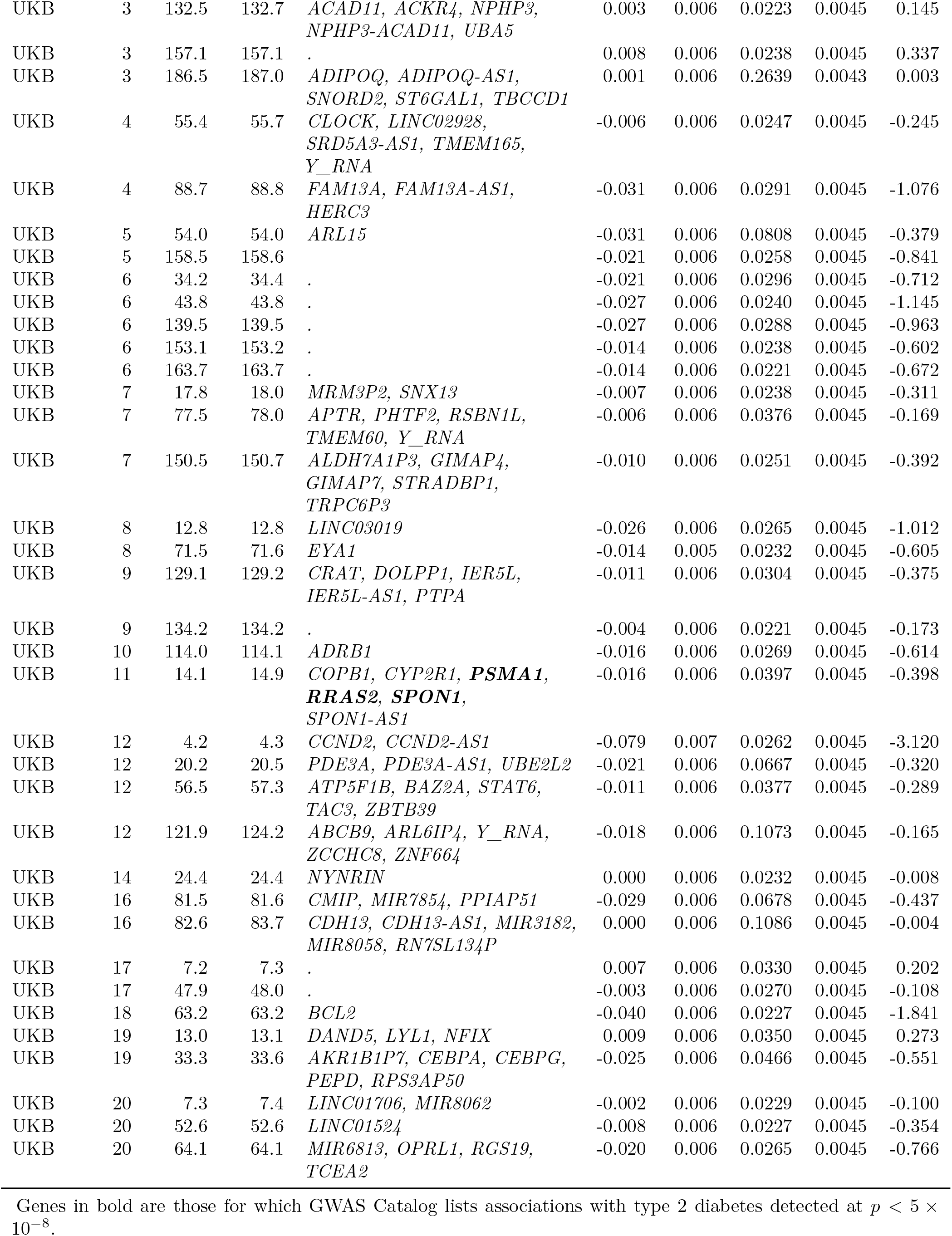
Summary statistics for regression of type 2 diabetes and adiponectin levels on genetic instruments.

